# PANDORA: An AI model for the automatic extraction of clinical unstructured data and clinical risk score implementation

**DOI:** 10.1101/2024.09.18.24313915

**Authors:** Natalia Castano-Villegas, Isabella Llano, Daniel Jimenez, Julian Martinez, Laura Ortiz, Laura Velasquez, Jose Zea

## Abstract

**Introduction:** Medical records and physician notes often contain valuable information not organized in tabular form and usually require extensive manual processes to extract and structure. Large Language Models (LLMs) have shown remarkable abilities to understand, reason, and retrieve information from unstructured data sources (such as plain text), presenting the opportunity to transform clinical data into accessible information for clinical or research purposes.

**Objective:** We present PANDORA, an AI system comprising two LLMs that can extract data and use it with risk calculators and prediction models for clinical recommendations as the final output.

**Methods:** This study evaluates the model’s ability to extract clinical features from actual clinical discharge notes from the MIMIC database and synthetically generated outpatient clinical charts. We use the PUMA calculator for Chronic Obstructive Pulmonary Disease (COPD) case finding, which interacts with the model and the retrieved information to produce a score and classify patients who would benefit from further spirometry testing based on the 7 items from the PUMA scale.

**Results:** The extraction capabilities of our model are excellent, with an accuracy of 100% when using the MIMIC database and 99% for synthetic cases. The ability to interact with the PUMA scale and assign the appropriate score was optimal, with an accuracy of 94% for both databases. The final output is the recommendation regarding the risk of a patient suffering from COPD, classified as positive according to the threshold validated for the PUMA scale of equal to or higher than 5 points. Sensitivity was 86% for MIMIC and 100% for synthetic cases.

**Conclusion:** LLMs have been successfully used to extract information in some cases, and there are descriptions of how they can recommend an outcome based on the researcher’s instructions. However, to the best of our knowledge, this is the first model which successfully extracts information based on clinical scores or questionnaires made and validated by expert humans from plain, non-tabular data and provides a recommendation mixing all these capabilities, using not only knowledge that already exists but making it available to be explored in light of the highest quality evidence in several medical fields.

## INTRODUCTION

One of the most important sources of bias in observational research is the quality of the secondary information sources, often clinical registries [1,2]. Investigators and physicians are met with the fact that 80% of clinical data is unstructured [3]or only partially structured. Thousands of pieces of valuable information are buried in text formats in physicians’ notes or lost in low-quality databases with high percentages of missing data, unlabeled information, calculation errors, and defective formatting, just to name a few [4].

This poor data structuring may seem harmless when looking at individual cases. Although time-consuming, researchers and research assistants can “dig” the information out of other sources, such as clinical records, laboratory results, notes from treating physicians, and patients themselves, as a last resort.

Advances in technology and computational sciences have led to the ability to collect, organise, operate, analyse, and interpret vast amounts of data (i.e., Big Data)[5–7] and to program computers to reproduce repetitive, time-consuming tasks only performed by humans in past decades (i.e., Machine Learning or ML) [8–10]. It would be only logical to take advantage of these advances and use them to improve the way health systems work in terms of faster diagnosis, personalized risk management, accurate classification of disease, and fairer resource distribution. In this line of thought, when we analyze the impact of unstructured data, understanding that it is one of the most prominent sources of information to create solutions with Artificial Intelligence (AI), poor structuring is not only harmful on an individual scale but could also introduce bias in worldwide used algorithms that could affect millions of patients, their families and have a significant impact on the economy [11–14].

The first clinical registries digitalized in the 1960s were simple electronic records of specific patient notes. It was not until the 1990s that the digitalization of healthcare became more popular in light of computational development, which demanded more effective ways of handling information. However, it was the new millennium that brought about the widespread institution of clinical digitalization [15– 19]. Undoubtedly, it made it easier for clinicians and researchers to access and collect the patient’s medical data.

Nevertheless, the large flow of daily information, the short time a clinician has to evaluate a patient, and the overload of clinics, hospitals, and outpatient services curtail the quality of registered information. Also, the limited funding for architectural and technological infrastructure in health care contributes to the fact that the wide implementation of these technologies is often the exception rather than the rule [20–22].

In response to the need for a more straightforward, effective, and precise way to approach the issue of unstructured data in health systems and research, we sought to utilize the advances made in Natural Language Processing (NLP) to deliver an AI solution that would actively help health-care personnel to find any piece of information they might need from clinical records, whether it is for research, diagnosis, urgent care or chronic care, etcetera. With this in mind, we created PANDORA.

In Greek mythology, PANDORA means the all-gifted. Inspired by this concept, we developed a robust algorithm framework that retrieves data from plain text and makes it accessible. We also propose a model that applies scores and clinical practice guidelines to the information retrieved, incorporating this capability in PANDORA. In the following sections, we explain how PANDORA came to be and give an initial scope of what could be achieved with its implementation in healthcare scenarios.

## METHODOLOGY

This section will briefly explain the methods and resources, including technical functions. They are referenced and available for consultation at the respective websites cited.

### General description

To explain how PANDORA was developed, it is helpful to divide the process into two smaller sections. PANDORA is a modular algorithm. Each section consists of an algorithm, referred to as an agent. One is responsible for extracting information from the Electronic Health Records (EHRs) and constitutes the Extraction Phase. The other uses the knowledge in clinical guidelines and validated scores chosen and provided by human researchers to predict a specific outcome based on the factors recovered from the EHRs. These factors are also referred to as features or variables. The latter algorithm constitutes the Recommendation Phase of the model, and the clinical score we chose for this validation was the PUMA scale for opportunistic case finding of Chronic Obstructive Pulmonary Disease (COPD) [23].

The two agents work synchronically. This means that the information extracted from the Electronic Information Records (EHRs) follows the instructions of clinical guidelines or score systems previously selected by the researchers (the PUMA scale in this case) to recover the variables needed (e.g. in the scoring system) and create a specific knowledge base, which will then be used to make a prediction or recommendation on the outcome of interest. The latter constitutes an intermediate step, the scoring or punctuation phase. PANDORA’s workflow is depicted in Figure 1.

**Figure 1.**
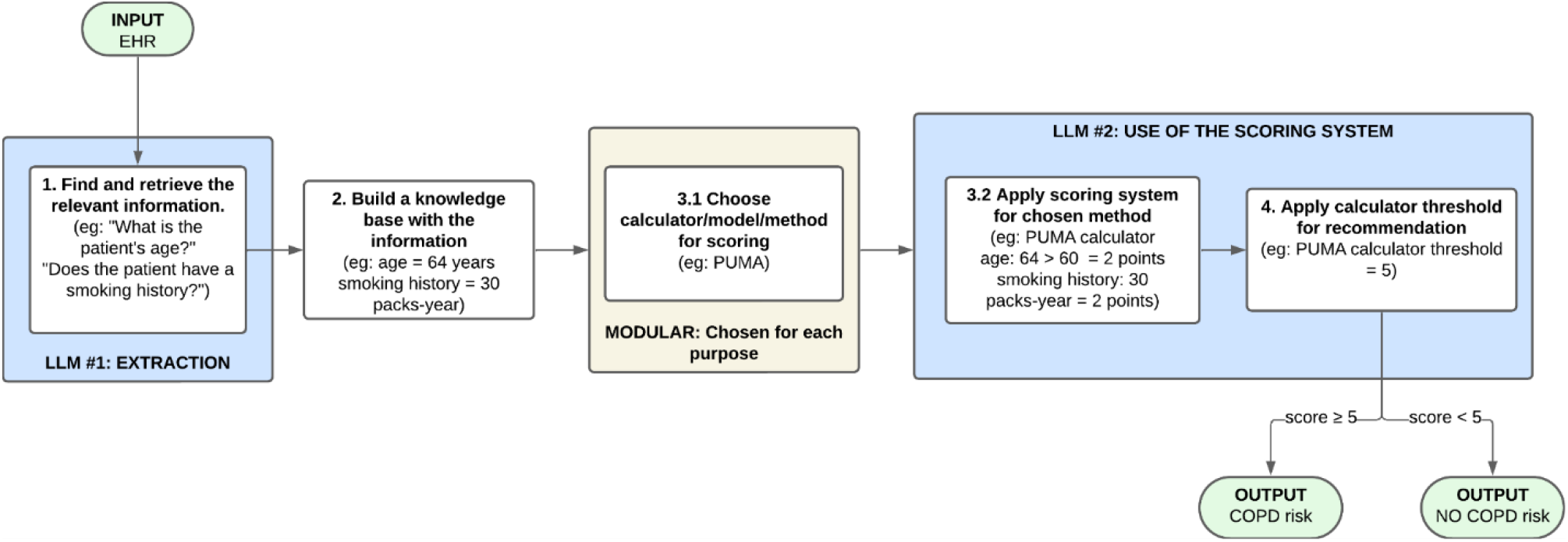
Workflow structure of PANDORA.

### General Sources of Data

To extract specific information from EHRs (Extraction Phase), we needed clinical cases or notes resembling real-life clinical charts’ structure. For this study we created two types of validations. The first used the Medical Information Mart for Intensive Care (MIMIC) database. This database contains data from previously deidentified Intensive Care Unit (ICU) patients hospitalised at the Beth Israel Deaconess Medical Center in Boston, USA. Its purpose is to assist quality research in healthcare and is available at https://mimic.mit.edu/. From 2002 to 2019, MIMIC collected patient data from two clinical information systems and has presented four updates. The latest is MIMIC-IV, where they added information from patients at the hospital and emergency department levels on top of the ones from previous versions at the ICU. Consequently, this version is divided into modules according to where the data was obtained from. One of these modules, the MIMIC-IV-Note, contains deidentified free-text clinical notes [24–26]. This was the database we used.

The second was a synthetic database generated automatically with an algorithm framework using the GPT family, following instructions provided by our medical team. They manually designed a standard form that simulated the structure of a clinical record made by the physician at a typical outpatient consultation in compliance with the Ministry of Health in Colombia [27].

Regarding the Recommendation Phase, we used Chronic Obstructive Pulmonary Disease (COPD) as the pathological entity of interest. We defined the presence or absence of risk for diagnosis of COPD as our primary outcome. This decision was based, first, on its high sub-diagnosis (89%) [28] and second, on the measurability of the outcome as a binary response that allowed us to evaluate the model’s overall reasoning after finding the relevant EHR information. Therefore, we used the standard clinical guideline for COPD, the Global Initiative for Chronic Obstructive Lung Disease (GOLD) 2024 Report, available at https://goldcopd.org/, and the PUMA COPD opportunistic case finding tool [23,29–31], including their latest validation study in 2022 [32].

The synchronism of both algorithms, which constitutes the scoring or punctuation phase of the model, did not require external input or further training, as it used the capabilities embedded into the PANDORA Large Language Model structure. The scoring was based on the mentioned scale (PUMA), which calculates a score for COPD risk employing seven features, namely sex, age in years, tobacco consumption in packets/year, dyspnea, chronic expectoration, chronic cough and whether the patient has had spirometry before. Each feature is assigned a score from zero to two, with a minimum result of zero and a maximum of nine, where risk is defined as a score more than or equal to five (Table 1.).

**Table 1.** The PUMA calculator.

| Variable | Sex |  | Age (years) |  |  | Smoking history (packs-year) |  |  | Dyspnea |  | Chronic cough |  | Chronic expectoration |  | Spirometry |  |
| --- | --- | --- | --- | --- | --- | --- | --- | --- | --- | --- | --- | --- | --- | --- | --- | --- |
|  | F | M | 40-49 | 50-59 | >60 | <20 | 20-30 | >30 | Yes | No | Yes | No | Yes | No | Yes | No |
| Score | 0 | 1 | 0 | 1 | 2 | 0 | 1 | 2 | 1 | 0 | 1 | 0 | 1 | 0 | 1 | 0 |

## Materials

All phases were developed using Python 3.12.2, Microsoft Office 365, the Arkangel App capabilities (https://www.arkangel.ai/), the AI translation and writing assistant Deepl (https://www.deepl.com), and the other cited resources from the internet.

### Algorithmic framework

PANDORA uses Natural Language Processing (NLP) algorithms and statistical algorithms to extract and analyse data from Electronic Health Records (EHRs). The primary algorithmic framework refers to the algorithms’ capabilities needed to perform at the different phases. It includes the following:

1. **Natural Language Processing (NLP)** techniques process and extract relevant disease-related factors from unstructured text within EHRs. This includes using models that understand medical terminology and context, allowing for accurate information extraction.
2. **Chain of Thought Strategy (CoT)**: This strategy ensures that the sequence of reasoning is maintained when extracting and analysing data. PANDORA can accurately map or associate patient data with disease factors by following a logical progression. CoT consists of reasoning steps, breaking down questions to guide language models through multi-step reasoning problems. This technique has been shown to improve LLM reasoning [33].
3. **Non-Relational Database Algorithms**: To manage the knowledge base, non-relational database algorithms efficiently store and retrieve patient-specific factors, allowing quick access during the recommendation process.
4. **Clinical algorithms:** Recommendations are constructed based on results from clinical algorithms. Here, we employ the PUMA calculator to screen for COPD risk.

### System development

Based on the previous framework, we develop a system with three main components:

1. **EHRs Data Extraction**: This stage involves extracting critical information related to disease factors from clinical records using advanced Natural Language Processing (NLP) techniques. The process leverages the chain of thought strategy to ensure that all relevant data is accurately identified and captured from unstructured text.
2. **Knowledge Base Construction**: The extracted information is then used to build a non-relational knowledge base. This knowledge base stores all pertinent patient data related to the identified disease factors using the features of a particular validated clinical guideline or score, serving as a structured repository that supports the next stage of the process.
3. **Recommendation System**: Instead of directly inferring from the guidelines, PANDORA employs a recommendation mechanism. The model takes the information stored in the knowledge base (from EHRs) and, following the predefined instructions (specific disease scores), suggests whether a patient is at risk of having a particular disorder. This recommendation is based on analysing the extracted factors and their alignment with established medical criteria or methodology. In this case, the recommendation comes from the PUMA calculator.

Once these steps are taken, PANDORA’s first assistant extracts the information, if available, from the EHRs and builds a knowledge base that is passed onto the second one, which runs the PUMA calculator on the extracted data and recommends whether to conduct further testing for COPD based on the presence or absence of risk, according to the score.

### Algorithm Evaluation

This score gives lower marks for incomplete answers or those with redundant information. It is calculated as the mean cosine similarity of the original question to a series of artificial questions that are reverse-engineered from the answer [36]. All metrics and their interpretations are tabulated in Table 2.

**Table 2:** Quality of text summarization metrics and their interpretation.

| Metric |  | Interpretation |
| --- | --- | --- |
| BERTScore |  | This score reflects the system's ability to evaluate the semantic similarity between the generated and reference texts, indicating high accuracy in understanding and generating relevant responses. |
| SemanticScore |  | This metric measures the semantic coherence of the output, demonstrating PANDORA's effectiveness in maintaining the meaning and context of extracted information. |
| RelevanceScore |  | This score indicates how well the extracted information aligns with the relevant disease factors, ensuring that the most pertinent data is used in the recommendation process. |
| Judge Alignment Metrics | Accuracy | These are the metrics PANDORA got when comparing its answers to another LLM's responses, in this case, Cloud AI. |
|  | Recall/Sensitivity |  |
|  | F1 Score |  |

### Evaluation of the Extraction Phase

To evaluate the model’s ability to extract information from the clinical notes, we used the EHR-DS-QA dataset found at https://physionet.org/content/ehr-ds-qa/1.0.0/. Its authors designed questions to evaluate LLMs’ extraction capabilities. They created this dataset with clinical questions and answers (QA pairs) using the LLM Meta Llama 2 for AI generation. The sources for the QA pairs were the clinical discharge notes from the MIMIC-IV-Note database mentioned above. It sampled 21466 medical discharge summaries from MIMIC-IV and automatically generated an outcome of 156,599 QA pairs. Based on convenience, we selected a subset of 506 from those original QA pairs since they correspond to cardiorespiratory clinical cases and had been reviewed by human physicians. This way, we had our reference standard for comparing PANDORA’s text extraction (Figure 2.).

**Figure 2.**
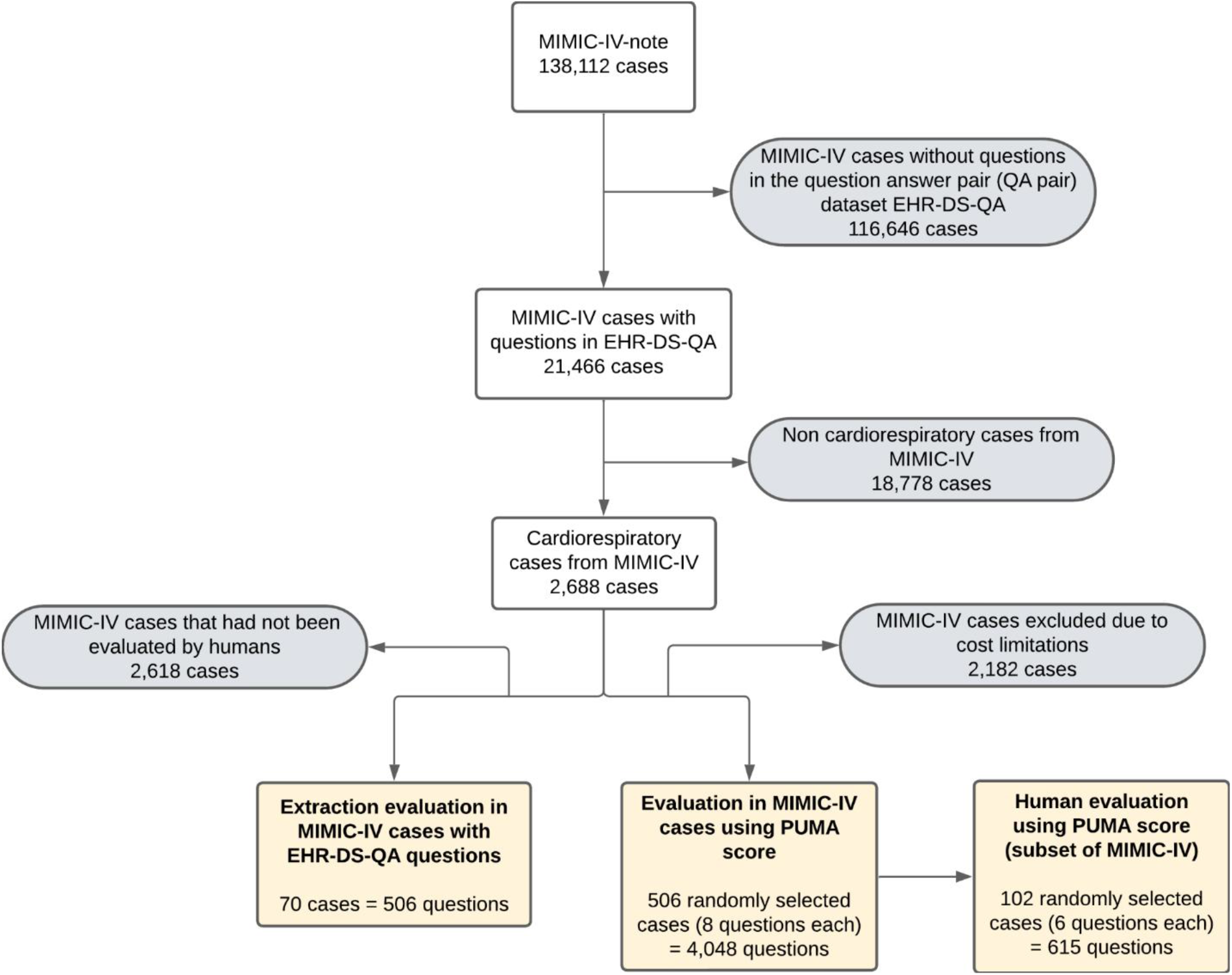
Flow chart of the MIMIC-IV-Note database and sample size.

An example of a question found in EHR-DS-QA is: “Does the patient have any known allergies or adverse drug reactions?” If the extraction was correct, the EHR should state the response [33]. No preprocessing was applied to the input data since the goal was to directly manage and analyze complex, unstructured data from EHRs without altering the original content.

Then, we employed four metrics to measure the quality of text summarization against the EHR-DS-QA dataset benchmark answers. Initially, we used BERTscore, an automatic metric used in text generation that calculates the similarity between a candidate and a reference sentence. The SimilarityScore is evaluated using contextual embeddings. This concept refers to how words are represented as vectors that algorithms in natural language processing can use to understand and produce language from them. It can recognize linguistic structures and their possible definitions instead of comparing exact matches between words [34]. The SemanticScore uses the same method as BERTscore but employs more semantic embeddings, better suited to understanding more subtle language characteristics, such as paraphrasing. RelevanceScore assesses the correctness and completeness of the answer [35].

Additionally, we assessed extraction using Judge Alignment Metrics. This strategy was developed as a more scalable and automated alternative to human evaluation [37]. It employs state-of-the-art LLMs like GPT-4o or Gemini to judge PANDORA’s wording of responses. In this case, we built a confusion matrix with the results and calculated accuracy, precision, recall, and F1 (Table 3.). The Judge LLM was given the exact information as PANDORA, including the EHRs, the PUMA score, and the GOLD 2024 guidelines. The LLM we used as judge was Cloud AI from *Devsig Technologies Private Limited*, which “is based on the Generative Pre-trained Transformer architecture and is pre-trained to generate human-like text” [38]. This application is free to access and available at https://play.google.com/store/apps/details?id=com.devsig.cloudai&hl=en&pli=1.

**Table 3.** Semantic scores for extraction evaluation using the EHR-DS-QA dataset.

| Metric |  | Score |
| --- | --- | --- |
| BERTScore |  | 0.911 |
| SemanticScore |  | 0.925 |
| RelevanceScore |  | 0.901 |
| Judge Alignment Metrics | Accuracy | 0.838 |
|  | Recall/Sensitivity | 0.838 |
|  | F1 Score | 0.912 |

### Recommendation Phase Evaluation

Once the model’s general extraction capabilities were assessed, two strategies were implemented to evaluate PANDORA’s output.

### First Strategy

A two-step evaluation was designed to determine the model’s specific extraction capability, implementing the PUMA score. We used 506 randomly selected QA pairs from the EHR-DS-QA dataset to test it. This was a different set of 506 from the one in the previous section, which we used for semantic scores and Judgement Alignment Metrics, but also based on the clinical discharge notes from MIMIC-IV-Note. The reason was that we wanted to explore the performance of our model with real clinical cases with original diagnoses from pathologies with or without cardiorespiratory components. We used the same number of questions to maintain the number of cases reviewed, adding that using the total 2688 Cardiorespiratory cases was extremely costly. However, since this set was not human evaluated, we performed a human evaluation of a subgroup of (102 clinical cases, 20%) to confirm, first, that the retrieved information was consistent with the information registered in the original medical notes from the EHR-DS-QA dataset and to evaluate how accurate the extraction of variables from EHRs was, following the instructions from the features in PUMA. Figure 7. indicates outcomes for this stage. Figure 3. depicts an example of the human evaluation process.

**Figure 3.**
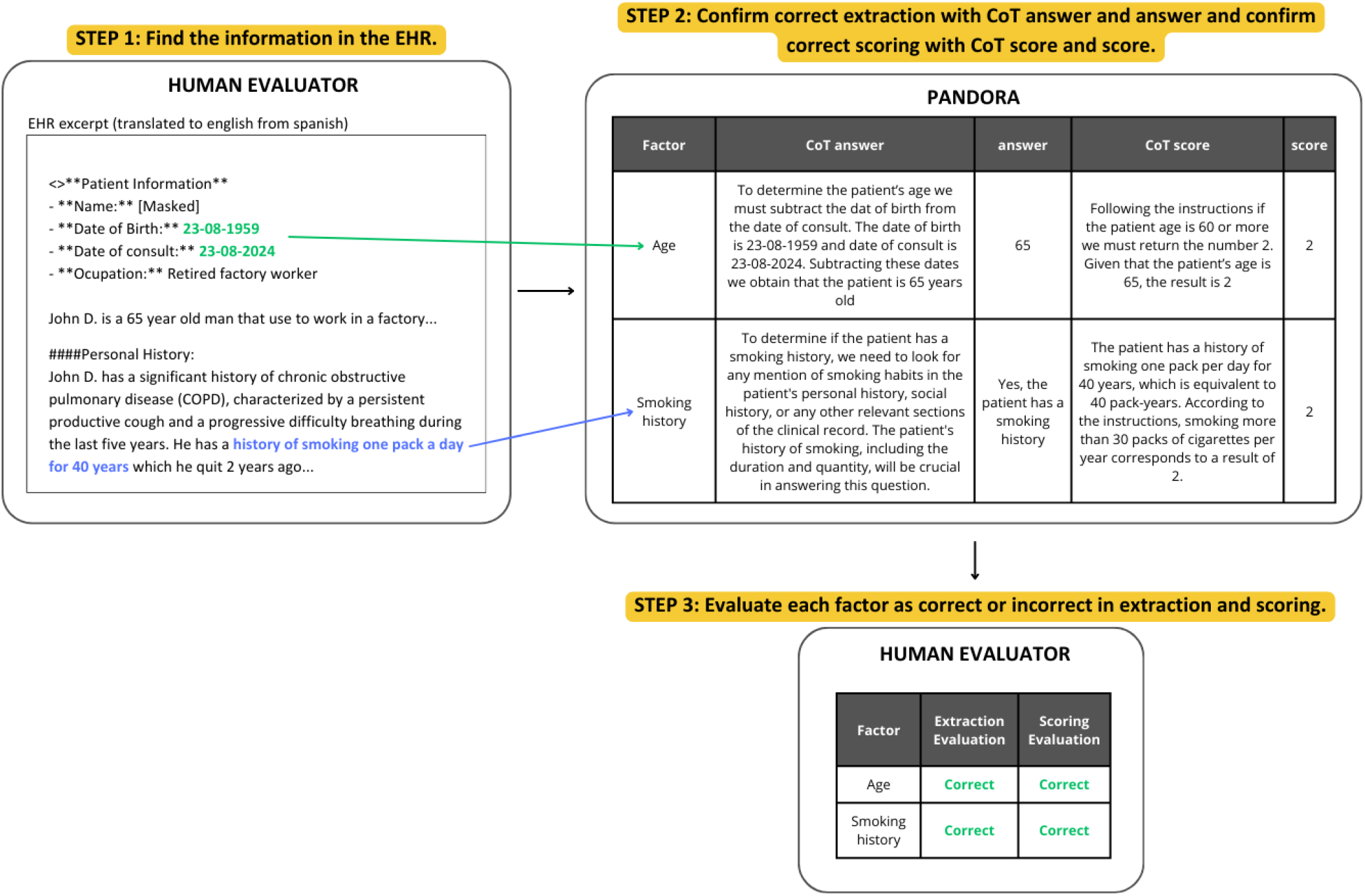
Examples of extraction and scoring capabilities assessment, performed in 3 steps by humans.

Second, we also assessed how the model made its recommendations using retrieved information. The diagnosis of COPD is defined as a ratio of Forced Expiratory Volume and Forced Vital Capacity (FEV1/FVC) lower than 0.70 in the first second after administering a dose of a bronchodilator in a pulmonary function test (the spirometry)[39,40]. The risk of diagnosis was exclusively assessed using the score of the Puma scale, which depends on 7 variables stated in Table 1. The maximum is 9 points; the minimum is 1 if male or 0 if a woman.

According to the validations made of the scale, in several countries in Latin America [23,28–31,41] and Asia [32], the optimal cutoff point is 5, as it was shown to detect more sub-clinical cases. For our study, we used the same threshold and defined that a score of 5 or more points on the PUMA scale would classify the individual as at risk for the diagnosis of COPD. However, this only represents the scoring capability of the model. To evaluate the diagnostic recommendation *per se*, PANDORA’s predictions were compared to the binary ground truth values: 1 (yes COPD risk) or 0 (no COPD risk). Ground truths are obtained from the EHRs in the original MIMIC-IV. The results are summarised in Table 4. by performance metrics of accuracy, sensitivity, specificity, and precision. The punctuation of the PUMA scale was evaluated as correct or incorrect and described using relative and absolute frequencies.

**Table 4.** Performance metrics for recommendation capabilities of PANDORA when using the MIMIC-IV database as standard, according to COPD’s previous diagnosis.

| Metric | Value |  |
| --- | --- | --- |
|  | Considering the history of COPD | Not considering the history of COPD |
| Sensitivity | 0.855 | 0.194 |
| Specificity | 0.700 | 0.925 |
| Precision | 0.815 | 0.800 |
| Accuracy | 0.794 | 0.480 |
| F1 score | 0.835 | 0.312 |
| Cohen's Kappa | 0.790 | 0.470 |

### Second Strategy

The other approach to evaluate the recommendation phase was the creation of synthetic clinical charts (synthetic EHRs) with a multi-step framework using the GPT family based on a guideline designed by our team’s medical lead. The document was built on the traditional structure of a Colombian-based outpatient consultation record and can be accessed in Supplementary Material 1.

By creating this synthetic database, we wanted to challenge PANDORA with clinical cases to test how accurately it could extract and apply the PUMA scale when the symptoms were similar to the extracted data. We used the nine possible differential diagnoses of COPD listed in Figure 5. Then, we manually created nine clinical cases using the said guideline, generated clinical records for each differential diagnosis and gave them as few-shot examples to the algorithmic framework for the elaboration of 100 synthetic clinical cases. Some of the instructions given to PANDORA (action also known as prompting) during this stage can be found in Figure 4.

**Figure 4.**
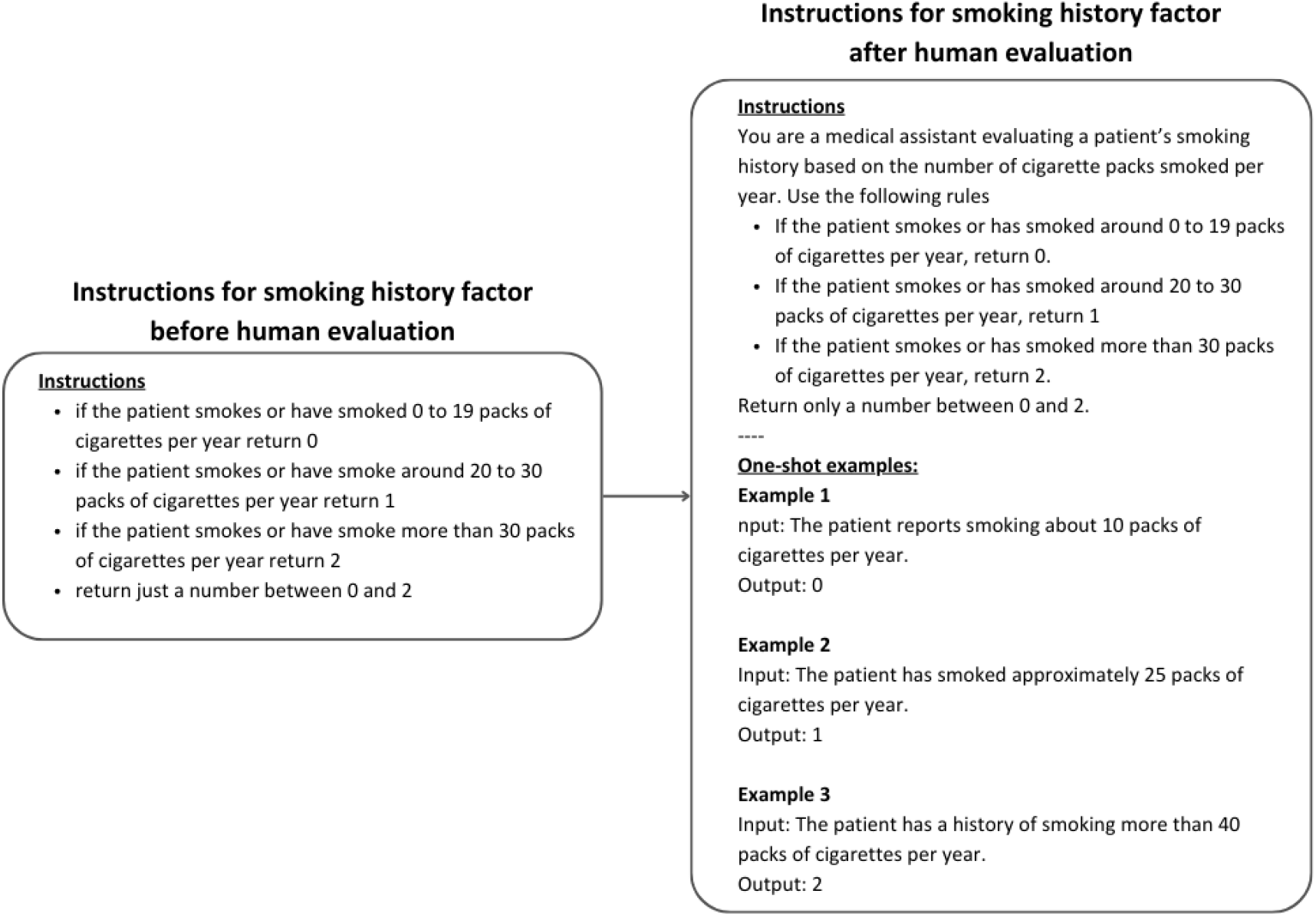
Example of an instruction or prompt given to PANDORA on how to score the individual’s “smoking history” based on the PUMA scale. Original instruction (left) and improved instruction after review (right)

**Figure 5.**
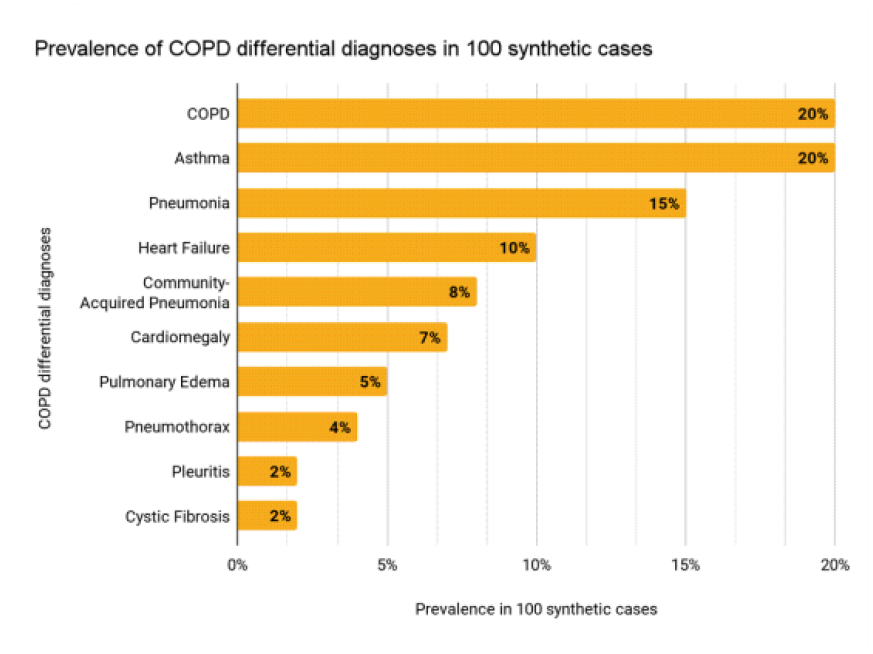
Differential diagnoses of COPD used to simulate outpatient consultation records.

At each process step, our medical and Machine Learning (ML) teams worked together to improve prompting strategies and recreate the outpatient scenario as closely as possible. Supplementary Material 2 provides an example of a synthetic clinical case. Similarly to the evaluation performed in the first strategy described above, we evaluated extraction, recommendation, and PUMA scale punctuation capabilities using relative and absolute frequencies, confusion matrices, and performance metrics.

Several EHRs stated a history of COPD. Therefore, we decided to introduce the presence of already diagnosed COPD as a feature to be extracted. If a patient had a previous history of COPD, the model was instructed to classify them as COPD-risk independent of their score in the PUMA. This strategy was only applied to the evaluations in MIMIC. The performance metrics for recommendation in these cases are in Table 4.

## RESULTS

This section will elaborate on PANDORA’s assessment results compared to the clinical discharge notes on the MIMIC-IV database and the synthetic outpatient clinical scenarios.

### Semantic Scores

The previous section abundantly described the meaning of these metrics (see Table 2). Table 3 demonstrates their results.

### Extraction, scoring and recommendation capabilities evaluation on the MIMIC-IV database

Based on the original 506 cases from the MIMIC-IV-Note and derived EHR-DS-QA database, we randomly chose a subset of 20% to evaluate them using human revisors. Figure 6. shows the results obtained from the human evaluation of the extraction and scoring capabilities of this subset of 102 cases. The variables age and smoking history had to be excluded in all cases because they were erased in the de-identification process of clinical records in the original MIMIC database and were unavailable. This resulted in 615 questions used to assess the scoring capability. Adequate extraction was demonstrated in 100% of questions, while 581 (94.47%) were classified as presenting correct scoring under the PUMA scale. Most of the 34 mistakes in scoring occurred when the model would not recognize COPD cases if the diagnosis were already in the clinical chart as a past disease. Table 4 compares how the performance metrics for the recommendation capability changed when instructing PANDORA to classify an individual as at risk for COPD in the presence of a history of COPD diagnosis against only using a PUMA score ≥5. Sensitivity increased by 66% when a search for a previous diagnosis of COPD was included.

**Figure 6.**
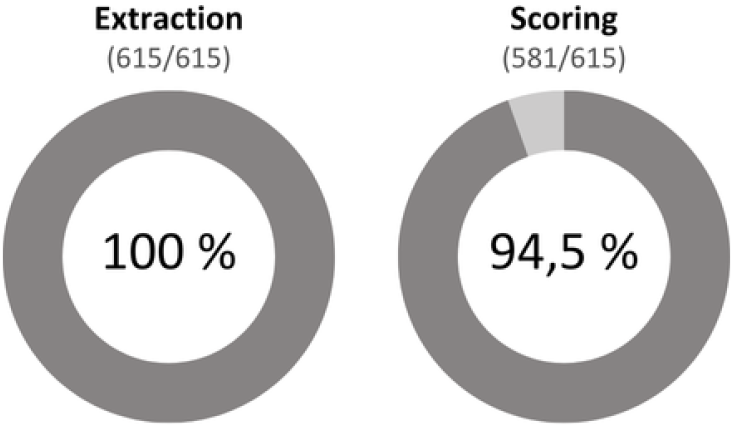
Human evaluation of extraction and scoring capabilities using the MIMIC-IV database as standard and based on the PUMA scale. The results express the accuracy of the test as the proportion of correct answers on each capability.

**Figure 7.**
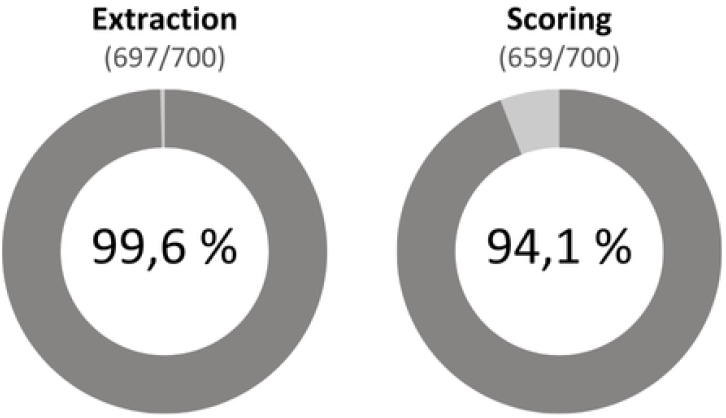
Human evaluation of extraction and scoring capabilities using the synthetic cases as standard and based on the PUMA scale. The results express the accuracy of the test as the proportion of correct answers on each capability.

Conversely, specificity decreased by 22.5%. Metrics were obtained from confusion matrices found in supplementary materials 3 and 4. On two occasions, the model hallucinated when it could not find data regarding the time frame that a specific symptom had been present and used similar reasoning (chain of thought) to classify its nature as acute or chronic.

### Extraction, scoring and recommendation capabilities evaluation on synthetic cases

The human evaluation of the 100 synthetic cases revealed optimal information extraction and scoring capabilities. As shown in Figure 7, PANDORA correctly extracted the information in 99.6% of cases. This means it demonstrated 697 adequate extractions from 700 questions, 7 for each feature in PUMA, applied to each synthetic clinical case and assigned the correct score, following the Puma scale in 94% of cases (658/700).

Two of the three extraction mistakes are associated with the assumption that having a history of COPD in the clinical record means the patient’s expectoration must be chronic. The other mistake occurs when PANDORA fails to recognize the phrase “worsening of the symptoms over the past few months” signifying their chronic nature.

Of the 41 scoring mistakes, 28 correspond to a misunderstanding from our LLM of the threshold values for the 3 categories of smoking history in PUMA, resulting in wrong punctuations for the risk of COPD. Of the remaining 13 scoring errors, three are due to an incorrect classification of age where PANDORA correctly calculated the patient’s age but still classified it into a mistaken age interval. The other 10 cases are associated with the definition of chronic cough and expectoration, where the model understands the term “persistent” as always referring to the chronic nature of symptoms or gets confused when it is not given a specific period for the duration of a given symptom.

Based on the extraction and scoring, PANDORA recommends risk for COPD (PUMA score ≥ 5) or “other” for any other respiratory disease. Table 5 shows the confusion matrix and performance metrics for recommendation compared to the original diagnosis in the synthetic case. PANDORA reached a sensitivity of 100% and a specificity of 20% since it incorrectly classified 64 synthetic cases at risk for COPD. Table 6. presents the distribution of PUMA in the false positive cases, stating their specific diagnosis and characteristics distribution.

**Table 5:** Confusion matrix and performance metrics for recommendation capabilities of PANDORA when using the synthetic cases as standard.

|  | Diagnosis COPD | Diagnosis Other |  |
| --- | --- | --- | --- |
| PANDORA COPD | 20 | 64 | 84 |
| PANDORA Other | 0 | 16 | 16 |
|  | 20 | 80 | 100 |

| Metric | Value |
| --- | --- |
| Sensitivity | 1.0 |
| Specificity | 0.200 |
| Precision | 0.238 |
| Accuracy | 0.360 |
| F1 score | 0.385 |
| Cohen's Kappa | 0.347 |

**Table 6:** Absolute frequencies of PUMA features in the misdiagnosed individuals, organized by differential diagnosis.

| Diagnosis |  | Sex<br>(M:F) | Age |  |  |  |  |  | Smoking history (packs/year) |  |  |  |  |  | Dyspnea |  | Chronic<br>Cough |  | Chronic<br>Expectora<br>tion |  | Spirometry |  |
| --- | --- | --- | --- | --- | --- | --- | --- | --- | --- | --- | --- | --- | --- | --- | --- | --- | --- | --- | --- | --- | --- | --- |
|  |  |  | 40-49 |  | 50-59 |  | >60 |  | <20 |  | 20-30 |  | >30 |  |  |  |  |  |  |  |  |  |
| COPD differential<br>diagnosis | n |  | (n) | (%) | (n) | (%) | (n) | (%) | (n) | (%) | (n) | (%) | (n) | (%) | (n) | (%) | (n) | (%) | (n) | (%) |  |  |
| Asthma | 17 | 13:4 | 0 | 0 | 10 | 59 | 7 | 41 | 16 | 94 | 0 | 0 | 1 | 6 | 17 | 100 | 17 | 100 | 1 | 6 | 17 | 100 |
| Pneumonia | 12 | 11:1 | 0 | 0 | 0 | 0 | 12 | 100 | 0 | 0 | 0 | 0 | 12 | 100 | 12 | 100 | 1 | 8 | 3 | 25 | 0 | 0 |
| Heart Failure | 8 | 8:0 | 0 | 0 | 0 | 0 | 8 | 100 | 2 | 25 | 1 | 13 | 5 | 63 | 8 | 100 | 7 | 88 | 0 | 0 | 8 | 100 |
| Cardiomegaly | 6 | 6:0 | 0 | 0 | 0 | 0 | 6 | 100 | 0 | 0 | 0 | 0 | 6 | 100 | 6 | 100 | 3 | 50 | 0 | 0 | 0 | 0 |
| Community Acquired-Pneumonia | 3 | 3:0 | 0 | 0 | 0 | 0 | 3 | 100 | 0 | 0 | 0 | 0 | 3 | 100 | 3 | 100 | 0 | 0 | 0 | 0 | 0 | 0 |
| Pulmonary Edema | 5 | 5:0 | 0 | 0 | 0 | 0 | 5 | 100 | 1 | 20 | 0 | 0 | 4 | 80 | 5 | 100 | 0 | 0 | 2 | 40 | 0 | 0 |
| Pulmonary Fibrosis | 8 | 8:0 | 0 | 0 | 0 | 0 | 8 | 100 | 0 | 0 | 0 | 0 | 8 | 100 | 8 | 100 | 8 | 100 | 0 | 0 | 7 | 88 |
| Cystic Fibrosis | 1 | 1:0 | 1 | 100 | 0 | 0 | 0 | 0 | 1 | 100 | 0 | 0 | 0 | 0 | 1 | 100 | 1 | 100 | 1 | 100 | 1 | 100 |
| Pneumothorax | 4 | 4:0 | 0 | 0 | 1 | 25 | 3 | 75 | 0 | 0 | 0 | 0 | 2 | 50 | 4 | 100 | 0 | 0 | 0 | 0 | 0 | 0 |
| Total | 64 | 59:5 | 1 | 2 | 11 | 17 | 52 | 81 | 20 | 31 | 1 | 2 | 41 | 64 | 64 | 100 | 37 | 58 | 7 | 11 | 33 | 52 |

Of the wrongly classified individuals, 92.2% were male, 81.25% were older than 60 years old, 64.06% had a smoking history of over 30 packages per year, and 100% had dyspnea. Chronic cough (57.81%), chronic expectoration (10.94%) and spirometry (51.56%) are also present but more varied across diseases.

## DISCUSSION

Despite the gigantic advances in health, data science, and machine learning, most clinical data is still unstructured, which means it is not organised in databases, to facilitate their processing and interpretation. Consequently, health-related data that could be used to understand disease patterns and make high-impact decisions in public and private health systems is currently buried in rudimentary clinical software in plain text [42]. With our Generative AI PANDORA, we intend to provide a tool for the health industry that enhances the use of all the existing knowledge that has not yet been exploited.

Specifically, earlier or missed diagnoses of several diseases could be achieved by combining the vast amount of capabilities developed over the last couple of years in Natural Language Processing [43] and the traditional, validated clinical diagnostic scores and updated clinical guidelines that contain the best available evidence and the consensus of field experts around the world.

### Context

The literature has explored LLMs’ ability to extract information from various text sources, such as EHRs and clinical notes. However, most models offer information retrieval and recommendation functions separately [44–52], while others are machine learning algorithms focused on diagnosis and risk prediction from organised, tabular data [50,52–58].

To name a few examples, Gu et al. tested the capability of information extraction from free electronic health records of 5 open-source LLMs. They evaluated their ability to extract social determinants of health and calculated accuracy as the number of true extractions over the total number of questions. Maximum performance was achieved by openchat_3.5, with an accuracy of over 80% [53]. Wang et al. also evaluated the impact of implementing LLMs for data extraction compared to human evaluation in China. The researchers found that the AI-assisted process improved efficiency by 80.7%, significantly reducing human labour time. Nevertheless, the accuracy of manual entry was 99.08%. The study also reported that the model made mistakes in understanding Chinese clinical terminology [54].

A recent preprint by Wiest et al. presents LLM-AIx for information extraction from unstructured medical text. In this study, the researchers present an adaptable pipeline for information extraction using Llama-3 70B. The model has been used for several purposes: extraction of signs and symptoms to make a diagnosis, recovery of information for research, detection of the risk of suicide and anonymization of the clinical data. This last function has been tested and improved, reporting 99.9% specificity and 100% sensitivity. They also report extraction from The Cancer Genome Atlas (TCGA) dataset https://www.cancer.gov/ccg/about. It retrieved variables such as the number of lymph nodes examined, if they were positive for cancer cells and whether the resection margin was tumour-free, with an overall accuracy of 87% [55].

On the other hand, Yu-Tzu Lee explored the integration of the extraction capability and recommendation in his thesis paper, which is available as a preprint at https://arxiv.org/abs/2407.10453. He intended to test the enhancement of medication recommendations using LLMs to extract information from free-text notes. The study used the MIMIC-III and CYCH datasets, including diagnoses and medication histories, and tested 7 different LLMs. The study shows that one of the seven models (G-BERT) improves its performance when text information extracted by the LLM is added alongside the medication codes, going from an Area Under the Precision Recall Curve (AUPRC) of 76.75% to 77.6% [56]. A different approach to retrieval and recommendation was proposed by Ozan Unlu et al. [57], who developed a model that would retrieve information from EHRs according to predefined selection criteria to select appropriate candidates for the clinical study Co-Operative Program for Implementation of Optimal Therapy in Heart Failure (COPILOT-HF; ClinicalTrials.gov number, NCT05734690). Here, their assistant, named RECTIFIER, extracted information according to inclusion criteria and used exclusion criteria to recommend final candidates. This study’s sample selection, compared with revision from non-licenced study staff, had 92% sensitivity and 94% specificity.

To our knowledge, no other health-related LLMs are pursuing this dual objective. In particular, there is no description in the current literature of a modular LLM capable of producing information recovery as well as a diagnosis or stratification using that recovered information based on human-made clinical algorithms. Therefore, the research in this manuscript is our initial approach to validating a model that could simultaneously serve several purposes: information retrieval, integration with clinical guidelines, and recommendations for the risk of a diagnosis.

### Analyses and Findings

Our model demonstrated high-quality text summarization as evidenced by the BERTScore, which means that PANDORA can understand and generate relevant responses. Also, the Semantic Score demonstrates its effectiveness in maintaining meaning and context, and the Relevance Score indicates how well the information aligns with the relevant disease factors, showing that the model uses pertinent data in its recommendations. A Judge Alignment Metric was also applied; the good marks imply that our model performs well semantically in the presence of state-of-the-art LLMs and all their capabilities for evaluating the input (e.g., information retrieved from EHRs) and the output (recommendation regarding risk of diagnosis). Notwithstanding, these scores measure only one dimension of the written answer.

State-of-the-art (SotA) LLMs refer to the Large Language Models with the highest accuracies reported compared to other current LLMs [58]. For example, the current SotA performance for GPT-4 is 90.2% and 85.4% for Med-PaLM 2 when assessed in light of one of the reference standards. These reference standards mainly consist of enormous datasets of questions and answers built from medical board exam questions or telemedicine interactions [34,59,59–61], which provide revised answers and facilitate quantitative analyses. The strategies usually consist of running these databases on the LLM and comparing their responses to said standards [46,51,62–69]. Usually, the expected outcome is a binary classification of correct or incorrect, one or zero, etcetera, according to the nature of the standard dataset. From there, they calculate the accuracy, often the measure presented in the manuscripts [33,60,70], defined as the correct answers obtained with the model being tested divided by the total of items in the database.

Although no scientific consensus exists for a test that could be used as a gold standard in LLM evaluation [71], human expert revisors are still considered the desirable comparison criteria for LLM responses [72,73]. The Judge Alignment Metrics strategy (e.i. Using a benchmark LLM to evaluate another original LLM) was developed as a more scalable and automated alternative to human evaluation [37]. The rationale is that human eval, although ideal, is almost impossible given the sizes of clinical notes databases, some containing more than fifteen thousand questions or clinical scenarios. It would transform any attempt at developing an LLM into a costly, time-consuming matter and exhausting for the professionals involved.

The MIMIC-IV-Note database was explicitly chosen to test PANDORA for its comprehensive coverage of patient health records. This database focuses exclusively on healthcare, making it an ideal source for extracting disease-related factors. Furthermore, MIMIC-IV-Note has been widely used as a benchmark for training and evaluating models in the medical field [24–26]. As such, we used it as the standard of reference for the qualitative assessment of PANDORA. This assured the model’s exposure to raw, intricate data typical in clinical settings, allowing it to work effectively in this context. Also, we emphasised handling open-ended responses so the algorithms could extract relevant factors. Unlike other databases, there are no described benchmarks for LLM evaluation specifically using MIMIC IV.

PANDORA showed that it can adequately extract structured data from unstructured sources, such as medical records and discharge notes. To validate this, we performed Human Evaluations of each process step using a subset of the MIMIC-IV database. In this initial evaluation, our model demonstrated perfect extraction capability (100%); additionally, we explored its ability to interact with a validated risk calculator (scoring capability), the PUMA scale for COPD risk assessment, which revealed that our model understands the rationale behind the scoring rules.

Regarding the recommendation capability, it could point out the risk for COPD in all synthetic clinical cases and 89% of the MIMIC-IV discharge notes. Still, the specificity for this last capability was 20% for synthetic cases and 70% using MIMIC-IV. These results could be explained by the use of a highly sensitive COPD case-finding tool such as PUMA.

The “Prevalence and Usual Practice in a Population at-risk of COPD in General Medicine Practice in 4 Countries of Latin America” Study or PUMA [31] is described as an opportunistic case-finding tool for COPD [31], validated for screening in adult, heavy-smoker population. Thus, its threshold for COPD (≥ 5) risk was set accordingly. This raises the question of the tool’s applicability in a population with different baseline characteristics and risk factors: Could a different threshold be used? The scale’s precision is extensively described elsewhere [29,30,32,41] and is not the focus of this paper. However, it is plausible that setting a higher threshold could validate PUMA for use in a broader population base without pre-selected risk factors. Supplementary material 5. depicts the operative characteristics of the PUMA scale when applied to our population sample, using thresholds from 1 to 9. The calculations for operative characteristics of the PUMA scale, in one of the original samples it was validated, were presented by Lopez Varela et al. in their 2016 paper “Development of a Simple Screening Tool for opportunistic COPD Case Finding in Primary Care in Latin America: The PUMA study” [23].

However, both sources of clinical cases (MIMIC and synthetic) were tested using the same scale, which does not explain the 50% difference in specificity (70% for MIMIC, 20% for synthetic cases). We believe the explanation lies in the intrinsic characteristics of the sources used and our final aim.

The MIMIC-IV was a real-world dataset with patients who were entered into the ICU and had just been discharged. Their entire clinical record was that of a sicker patient. Unfortunately, we could not evaluate smoking habits or age, as it was part of the erased information in the de-identification process, mandatory for the public access of sensitive information. This fact makes the score results not comparable to those from the synthetic cases, which have all the information.

On the other hand, the synthetic cases were created to intentionally present the scale with cases that showed similar symptoms to COPD. The profile of this patient was also completely different, as they were outpatient consultations, healthier in general than the previous ones. For this database, the number of cases of PANDORA misclassified as COPD was 64, which shows the adequate extraction and scoring of the model but the high sensitivity and low specificity of PUMA. To elaborate on this, take the example of a 62-year-old man (male 1 pt+age 2 pts) presenting with dyspnea (1 pt) due to heart failure, with spirometry performed (1pt); this patient will be undoubtedly be classified as a case for COPD if only evaluated using the PUMA scale.

To approach this, we found out there were 62 synthetic cases that were given COPD as part of past clinical history by the generative model and decided to use this criterion for a subsequent analysis on MIMIC (presented in the results section) adding an item to the extraction phase, which we prompted as “*does the patient has a smoking history*?”. This approach improved sensitivity by 66%.

A remarkable contribution of the present research is the application of the recently proposed “Self-Thought Evaluators”, described in a paper by the same name, published on August 8, 2024 [74]. The approach proposes the evaluation of models without human judges. Instead, it presents an approach using only synthetic data with which an AI model evaluates the performance of another. Our experience developing and using the Judge Alignment Metric was remarkable in that it allows any kind of assessment of any number of entries or queries, facilitating and expediting analyses, comparisons and improvements. The downside is that AI judges could make mistakes in their judgement and would amplify possible biases introduced when they were created.

### Biases

First, regarding the benchmark used (MIMIC-IV/EHR-DS-QA), we could not assess the complete set of questions for each case using the PUMA scale since age and smoking history were missing in all cases. Thus, we were not able to control this from the data source. Consequently, we performed a human evaluation of each case and question-answer pairs, not necessarily to apply the score, which would undoubtedly affect the recommendation, but to ensure every other capability was conserved. As mentioned, human evaluation is the standard of reference desired but is a cumbersome task. The ideal way to approach this is to manually revise the percentage of the cases, ideally 100 and no less than 50 cases [72].

Given that all data in PANDORA is generated and evaluated synthetically, the system may not fully represent the diversity of EHRs in the general population. This approach could introduce biases into the model, as the synthetic data might not capture the full range of variability in real-world clinical data. Furthermore, clinical note structure may vary by country or institution. We had medical professionals and government guidelines to replicate outpatient clinical cases. We explicitly instructed the LLM to consider all races, sexes, occupations, nationalities and social backgrounds to make the rules more egalitarian. Still, continuous human monitoring and re-evaluation are necessary to ensure and supervise PANDORA’s and other LLMs’ outcomes.

## CONCLUSION

This initial evaluation is the first step towards the validation and launch of a clinical and research tool that will allow the application of diagnostic scores from different diseases to information previously trapped in formats that made it inaccessible. Even institutions without structured databases will be able to use it and make the most of all the knowledge currently not being utilised, written in plain text.

## Data Availability

All data produced in the present study are available upon reasonable request to the authors

## CONTRIBUTIONS

NCV Medical epidemiologist:

Research, validation, manuscript writing and editing

IL Biomedical engineer:

Research, validation manuscript writing

DJ Machine learning engineer:

Methodology, software, validation

JM Machine learning engineer:

Methodology, project administration

LO Biomedical engineer:

Methodology, project administration

LV President and Co-founder:

Supervision

JZ CEO and Co-founder:

Supervision

