## Supplementary Materials for "PANDORA: An AI model for the automatic extraction of clinical unstructured data and clinical risk score implementation"

***Supplementary Material 1. A summarised version of Internal guidelines for constructing synthetic out-patient clinical records using GPT4o (translated from the original in Spanish, using DeppL AI translation and writing assistant). The original text is available upon request.***

Clinical record sections:

- Identification
- Relevant past history: personal, family, psychosocial, controlled substances, surgeries, epidemiological
- Current clinical complaint
  - Consultation motive in the patient’s words
  - Necessary characteristics of patient's evolution or presentation. Anything the patient has manifested that is relevant to the clinical case.
- System revision: Things referred by the patient, by systems, that are not directly related to the current disease
  - Ex. neurological, ENT, respiratory, cardiovascular, digestive… etc
- Physical examination: Objective evaluation by the doctor described by systems (head to toe)
  - Vitals, general state, neurological state, head and neck, thorax and lungs, cardiac and vascular, abdomen, genital and urinary, skin, extremities
- Laboratory and imaging results: New and/or old results.
- Clinical summary: Doctor’s analysis of the patient’s condition taking into account all of the evaluated information and all that was referred by the patient.
  - A small summary of subjective findings
  - Analysis of objective findings
  - Conclusion with diagnostic impressions
  - Recommendations based on findings and conclusions
  - Indications of how to use medication if relevant
  - Indications of complementary exams if relevant
- ICD-10: Include the International Disease Code.

***Supplementary Material 2. An example of a Synthetic Clinical Case was created using AI (translated from the original in Spanish using DeppL AI translation and writing assistant). Original text available upon request.***

<>**Patient Information** **Patient's Name:** Mr. John Smith

- Name:** Mr. John Smith

- Date of Birth:** March 15, 1959

- Date of Consultation:** August 23, 2024

- Occupation:** Retired School Teacher

Mr. John Smith spent over 35 years as a dedicated high school history teacher. He is well respected in his community for his engaging teaching style and commitment to the success of his students. Since retiring five years ago, he has enjoyed spending his time gardening, walking in his neighbourhood and participating in local community events.

\### Relevant Past History

\#### Personal History:

- **Hypertension:** Diagnosed 10 years ago, well controlled with lisinopril 10 mg daily.

- Seasonal Allergies:** Mild allergic rhinitis, managed with over-the-counter antihistamines as needed.

- Pneumonia:** Episode of community-acquired pneumonia 5 years ago, successfully treated with antibiotics.

- No history of diabetes, chronic respiratory disease (e.g., asthma, COPD), or cardiovascular disease other than hypertension.

\#### Family History:

- **Father:** Died at 78 years of age from myocardial infarction; had a history of hypertension and hyperlipidaemia.

- Mother:** Died at age 80 from complications of type 2 diabetes; also had hypertension.

- Siblings:** One brother, aged 70 years, with a history of hypertension and type 2 diabetes; one sister, aged 68 years, with no significant medical history.

- No known family history of cancer, stroke or chronic respiratory disease.

\#### Psychosocial History:

- **Marital Status:** Married, lives with wife in a suburban area.

- Occupation:** Retired school teacher.

- Lifestyle:** Generally active, enjoys gardening and walking in the neighborhood.

- Mental Health:** No history of depression, anxiety or other mental health disorders. Currently experiencing anxiety related to his illness and possible hospitalisation.

- Social Support:** Strong support system from his wife and close friends in the community.

- No history of smoking, alcohol or substance abuse.

\#### Controlled Substances:

- **No history of controlled substance use, abuse, or dependence.

\#### Surgical History:

- **Apendectomy:** Performed at age 25, uncomplicated.

- Cholecystectomy: **Performed at age 50 due to gallstones, uncomplicated.

- No history of other major surgeries or medical procedures.

\#### Epidemiological History:

- **Travel:** No recent travel, particularly to areas with endemic infectious diseases.

- Exposure:** Denies any known exposure to sick contacts or recent hospitalizations.

- Immunizations:** Up to date with COVID-19 vaccination; has not received current season's influenza vaccine.

- Living Conditions:** Resides in a suburban area with no known environmental hazards or exposure to industrial pollutants.

- Pets:** Owns a dog, with no known zoonotic diseases.

This comprehensive history provides a detailed overview of Mr. John Smith's medical, family, psychosocial, controlled substance, surgical and epidemiological history, which is crucial for accurate diagnosis and effective treatment planning.

\### Current Clinical Complaint

Reason for consultation:** ** ‘I have worsened my symptoms.

‘I have had a worsening cough, shortness of breath and fever for five days.’

**Necessary features of evolution/presentation:** **

Mr. John Smith reports that his cough started as dry but became productive with greenish sputum over the past two days. He has experienced chills, sweating and fever up to 102°F (38.9°C). He also mentions sharp, pleuritic chest pain on the right side, which worsens with deep breathing or coughing. His shortness of breath is rated 7/10 and is exacerbated by exertion.

Associated factors:** ** Mr. Smith notes

Mr Smith notes decreased appetite, unintentional weight loss, generalised myalgia and persistent headache. Despite taking acetaminophen, his fever and malaise have only marginally improved. He denies gastrointestinal symptoms, recent travel, sick contacts or hospitalisations.

\### Systems Review

**Neurological

- Generalised myalgia

- Persistent headache

- Increased lethargy

Respiratory:** ** Respiratory

- shortness of breath (7/10 in severity, exacerbated by exertion)

- Acute, pleuritic chest pain on the right side, intensified by deep breathing or coughing

Musculoskeletal:** ** Generalised myalgia

- Generalised myalgia

Digestive:** ** Decreased appetite

- Decreased appetite

- Unintentional weight loss during the last week

Skin and adnexa:** ** No specific complaints or observations

- No specific complaints or observations

Cardiovascular:** ** No specific complaints or observations

- No specific complaints or observations

**Genitourinary:** No specific complaints or observations noted

- No specific complaints or observations noted

Otolaryngological:** **Otorhinolaryngological:** No specific complaints or observations noted

- No specific complaints or observations

\### Physical Examination

\#### Vital Signs:

- Temperature: 101.8°F (38.8°C)

- Blood Pressure: 138/82 mmHg

- Heart Rate: 98 beats per minute

- Respiratory Rate: 22 breaths per minute

- Oxygen Saturation: 92% on room air.

\#### General Appearance:

- Mr. Smith appears fatigued and slightly dyspneic at rest. He is alert and oriented but appears uncomfortable and anxious. He is sitting upright and appears to be in mild distress due to his symptoms.

\#### Head and Neck:

- Head: Normocephalic, atraumatic.

- Eyes: Clear conjunctivae, anicteric sclerae. Pupils equal, round and reactive to light.

- Ears: Tympanic membranes bilaterally clear.

- Nose: Nasal mucosa slightly congested, without discharge.

- Throat: Oropharynx clear, without erythema or exudates.

- Neck: Flexible, without lymphadenopathy, without jugular venous distension.

\#### Chest and Lungs:

- Inspection: Increased work of breathing, with use of accessory muscles.

- Palpation: No tenderness to palpation over the chest wall.

- Percussion: Dullness on percussion over the right lower lung field.

- Auscultation: Decreased breath sounds in the right lower lung field with crackles and bronchial breath sounds. No wheezing.

\#### Cardiac and Vascular:

- Heart: regular rhythm, no murmurs, gallops or rubs.

- Peripheral Pulses: 2+ bilaterally in the radial, dorsalis dorsi and posterior tibial arteries.

- No peripheral oedema noted.

\#### Abdomen:

- Inspection: Abdomen flat, no visible distension.

- Auscultation: Bowel sounds present and normal in all quadrants.

- Palpation: Abdomen soft, non-painful, no organomegaly or masses detected.

- Percussion: Tympanic throughout, with no changeable dullness.

\#### Skin and adnexa:

- Skin: Warm and dry, no rashes or lesions observed.

- Nails: No acropaquia or cyanosis.

\#### Extremities:

- No joint swelling or deformities.

- Full range of motion in all extremities.

- Strength 5/5 in upper and lower extremities bilaterally.

\## Laboratory and Imaging Results

\### Laboratory Results:

- **Complete Blood Count (CBC):** **

- White Blood Cell Count (WBC): 14.5 x 10^3/µL (Reference Range: 4.0-11.0 x 10^3/µL)

- Haemoglobin (Hgb): 13.2 g/dL (Reference range: 13.5-17.5 g/dL)

- Hematocrit (Hct): 39.8% (Reference range: 38.8-50.0%)

- Platelet Count: 250 x 10^3/µL (Reference Range: 150-450 x 10^3/µL)

- Basic Metabolic Panel (BMP):** ** **Basic Metabolic Panel (BMP):** ** Sodium (Na): 138 mmol/L

- Sodium (Na): 138 mmol/L (Reference range: 135-145 mmol/L)

- Potassium (K): 4.2 mmol/L (Reference range: 3.5-5.0 mmol/L)

- Chloride (Cl): 101 mmol/L (Reference range 98-107 mmol/L)

- Bicarbonate (HCO3): 24 mmol/L (Reference range: 22-29 mmol/L)

- Blood Urea Nitrogen (BUN): 18 mg/dL (Reference range: 7-20 mg/dL)

- Creatinine: 1.0 mg/dL (Reference range: 0.6-1.2 mg/dL)

- Glucose: 98 mg/dL (Reference Range: 70-99 mg/dL)

- C-Reactive Protein (CRP):** 45 mg/L (Reference range: <10 mg/L)

- Procalcitonin:** 0.8 ng/mL (Reference range: <0.1 ng/mL)

- Liver Function Tests (LFTs):** ** Aspartate Aminotransferases:** 0.8 ng/mL (Reference range: <0.1 ng/mL)

- Aspartate Aminotransferase (AST): 30 U/L (Reference range: 10-40 U/L)

- Alanine Aminotransferase (ALT): 28 U/L (Reference range: 7-56 U/L)

- Alkaline Phosphatase (ALP): 85 U/L (Reference range: 44-147 U/L)

- Total Bilirubin: 0.8 mg/dL (Reference Range: 0.1-1.2 mg/dL)

- Arterial Blood Gasometry (ABG):** ** **PH: 7.45 (Reference range: 7.45)

- pH: 7.45 (Reference Range: 7.35-7.45)

- PaCO2: 35 mmHg (Reference range: 35-45 mmHg)

- PaO2: 75 mmHg (Reference range: 75-100 mmHg)

- HCO3: 24 mmol/L (Reference range: 22-26 mmol/L)

- O2 saturation: 94% (Reference range: 95-100%)

\### Imaging Results:

- **Chest X-Ray (CXR):** [Insert Date].

- Date: [Insert Date]

- Findings: Right lower lobe consolidation with air bronchograms, consistent with pneumonia. No evidence of pleural effusion or pneumothorax. No cardiomegaly noted.

- Computed Tomography (CT) of the Chest:** ** Date: [Insert Date].

- Date: [Insert Date]

- Findings: Patchy consolidation in the right lower lobe with surrounding ground-glass opacities. No significant lymphadenopathy. No evidence of pulmonary embolism. Mild bronchial wall thickening.

- Electrocardiogram (ECG):** ** Date: [Insert Date].

- Date: [Insert Date]

- Findings: Normal sinus rhythm, no acute ST-T segment changes, no evidence of ischemia or infarction.

\### Medical Orders

\#### Medications:

1. Amoxicillin-Clavulanate** **Amoxicillin-Clavulanate** 2.

- Route of Administration:** Oral

- Presentation:** Tablet

- Dosage:** 875 mg/125 mg

- Frequency:** Twice a day for 10 days

2. **Acetaminophen

- Route of Administration:** Oral

- Presentation:** Tablet

- Dosage:** 500 mg

- Frequency:** Every 6 hours as needed for fever and pain

3. **Albuterol

- Route of Administration:** Inhalation

- Presentation:** Metered-dose inhaler

- Dose:** 2 inhalations

- Frequency:** Every 4-6 hours as required for respiratory distress

4. **Lisinopril

- Route of Administration:** Oral

- Presentation:** Tablet

- Dosage:** 10 mg

- Frequency:** Once a day

\#### Complementary Tests:

1. **Chest X-ray **Chest X-ray

- Purpose:** To evaluate for pneumonia or other pulmonary pathology

- Frequency/Time: **Stat

2. Complete Blood Count (CBC)** **Purpose:** To assess pneumonia or other pulmonary pathology **Frequency/Time:** Stat 2.

- **

***Supplementary material 3. Confusion matrix for recommendation capability on a sample of 102 cases from MIMIC when previous COPD diagnosis was considered***

|  | **Diagnosis**  **COPD** | **Diagnosis**  **other** |  |
| --- | --- | --- | --- |
| **PANDORA**  **COPD** | 53 | 12 | 65 |
| **PANDORA**  **other** | 9 | 28 | 37 |
|  | 62 | 40 | 102 |

***Supplementary material 4. Confusion matrix for recommendation capability on a sample of 102 cases from MIMIC when previous COPD diagnosis was not considered***

|  | **Diagnosis**  **COPD** | **Diagnosis**  **other** |  |
| --- | --- | --- | --- |
| **PANDORA**  **COPD** | 12 | 3 | 15 |
| **PANDORA**  **other** | 50 | 37 | 87 |
|  | 62 | 40 | 102 |

***Supplementary Material 5. Performance metrics using cut-off points from 1-9 on the PUMA scale for assessing PANDORA in synthetic cases***

| **Metric** | **Cut-off point** | | | | | | | | |
| --- | --- | --- | --- | --- | --- | --- | --- | --- | --- |
|  | **>=1** | **>=2** | **>=3** | **>=4** | **>=5** | **>=6** | **>=7** | **>=8** | **=9** |
| **Sensitivity** | 1.0 | 1.0 | 1.0 | 1.0 | 1.0 | 1.0 | 1.0 | 1.0 | 0.700 |
| **Specificity** | 0.0 | 0.0 | 0.036 | 0.038 | 0.200 | 0.450 | 0.738 | 0.888 | 1.0 |
| **Precision** | 0.200 | 0.200 | 0.200 | 0.206 | 0.238 | 0.313 | 0.488 | 0.690 | 1.0 |
| **Accuracy** | 0.200 | 0.200 | 0.223 | 0.230 | 0.360 | 0.560 | 0.790 | 0.910 | 0.940 |
| **F1 score** | 0.333 | 0.333 | 0.333 | 0.342 | 0.385 | 0.476 | 0.656 | 0.816 | 0.824 |
| **Cohen's Kappa** | 0.184 | 0.184 | 0.208 | 0.214 | 0.347 | 0.551 | 0.786 | 0.908 | 0.939 |
